# Deep Learning Reconstruction versus Hybrid Iterative Reconstruction for Spectral CT: A Multi-Institutional Image Quality Assessment

**DOI:** 10.64898/2026.09.10.26362104

**Authors:** Matthew Nazarian, Amy E. Perkins, Eddy Y. Wong, Petra C. Koopmans, Merav B. Stromza, Leonid Roshkovan, Manoj Mathew, Jonathan Dorff, Jeffrey M Levsky, Matthew S. Lazarus, Peter B Noël, Leandro Slipczuk, Ali Dhanaliwala

**Affiliations:** Department of Radiology, Hospital of the University of Pennsylvania, Philadelphia, PA; Philips Healthcare, Orange Village, Ohio, USA; Department of Radiology, Montefiore Health System/Albert Einstein College of Medicine, New York, NY, USA; Division of Cardiology, Montefiore Health System/Albert Einstein College of Medicine, New York, NY, USA

## Abstract

**Purpose:** To compare image quality between deep learning reconstruction and hybrid iterative reconstruction for conventional and virtual monoenergetic images derived from the same spectral CT examinations.

**Methods and Materials:** This retrospective, post hoc secondary analysis of a multi-institutional, controlled, blinded reader study evaluated head, chest, cardiac, and abdomen/pelvis CT examinations acquired using dual-layer spectral CT. The parent study (clinicaltrials.gov identifier NCT07108205) included 147 examinations across a range of acquisition and reconstruction parameters. Raw data were reconstructed using hybrid iterative reconstruction (HIR; iDose^4^, Philips Healthcare) and spectral deep learning reconstruction (DLR; Spectral Precise Image, Philips Healthcare). Conventional images and corresponding virtual monoenergetic images (VMI) were generated with both methods from identical raw data. Randomized image pairs were independently evaluated by two blinded board-certified radiologists or cardiologists, and image quality was scored on a 5-point Likert scale. A post hoc analysis was performed on previously collected scores from a subset of examinations acquired at standard resolution and reconstructed with thin sections and a soft tissue kernel to focus comparison on HIR versus DLR. Least squares means were estimated using a model accounting for multiple readers per examination. Noninferiority of DLR to HIR was tested; when established, superiority was tested using the same model.

**Results:** 59 CT examinations from 56 patients met technical criteria for this analysis. DLR was noninferior and superior to HIR for head, abdomen, and cardiac examinations across conventional images and VMI, with p values ranging from <.0001 to .0058. DLR was noninferior but not superior for conventional chest images (p = .1430). Noninferiority was not established for chest VMI, although mean scores numerically favored DLR. This may reflect a ceiling effect from high HIR scores.

**Conclusion:** Spectral DLR produced image quality that matched or exceeded HIR across most anatomic regions and image types, including conventional and virtual monoenergetic images.

## Introduction

Image quality in computed tomography (CT) reflects a balance among radiation dose, image noise, and spatial resolution, such that an improvement in one is typically achieved at the expense of another.^1^ Managing this trade-off, delivering diagnostic image quality at the lowest reasonable dose, remains a central goal of CT image reconstruction.^2^

Iterative reconstruction (IR) reduces image noise relative to filtered backprojection and enables dose reduction, and hybrid IR (HIR) is now a clinical standard.^3^ However, at higher denoising strengths, HIR changes the appearance of the image, producing a blotchy or “plastic” texture and a shift in the noise power spectrum, and these changes can degrade low-contrast detectability even as the measured noise magnitude falls.^1,3^ Deep learning reconstruction (DLR) has advanced this goal further, offering greater noise reduction than HIR while better preserving noise texture and spatial resolution and supporting more aggressive dose reduction.^4-6^ These gains make DLR an attractive successor to HIR for clinical imaging. Early DLR implementations could, at high denoising strengths, over-smooth images and shift noise texture toward lower spatial frequencies, with potential effects on perceived spatial resolution and low-contrast detectability^7,8^. Preserving image texture while retaining the noise advantage has become a central aim of DLR development.^9^

These considerations are amplified in spectral CT. Dual-layer detector spectral CT generates conventional images together with spectral-derived images, virtual monoenergetic images (VMI), effective atomic number maps, and iodine density maps, from a single acquisition.^10,11^ Material decomposition tends to increase noise, and noise present in the raw data extends into the spectral-derived images and can degrade their quantitative accuracy.^12,13^ Iodine quantification on dual-layer detector CT is accurate, for example, second-generation dual-layer detectors quantify iodine within approximately ±0.5 mg/mL.^14^ Reducing raw-data noise is therefore expected to improve the visual quality of conventional images and the stability of the spectral-derived measurements, making a texture-preserving DLR attractive in this setting.

Much of the peer-reviewed evidence for this class of algorithm concerns conventional CT. Patient-derived phantom studies of the conventional algorithm have reported substantial dose-reduction potential without loss of image quality,^15^ and early clinical results have shown preserved image quality and lesion conspicuity at low dose.^16^ The reconstruction evaluated in the present study is the spectral implementation, a spectral deep learning reconstruction (spectral DLR; Spectral Precise Image, Philips Healthcare) that denoises both the conventional images and the spectral-derived images produced from the same acquisition (Figure 1). Direct evidence on this spectral implementation is more limited. In phantom work, dual-energy CT equipped with deep learning reconstruction has shown improved spectral performance, including reduced noise and better low-keV monoenergetic image quality,^17^ and a single-center clinical study rated DLR above iterative reconstruction.^18^ These reports are predominantly phantom-based or limited to single centers, and they have primarily characterized the quantitative performance of the algorithm.

**Figure 1.**
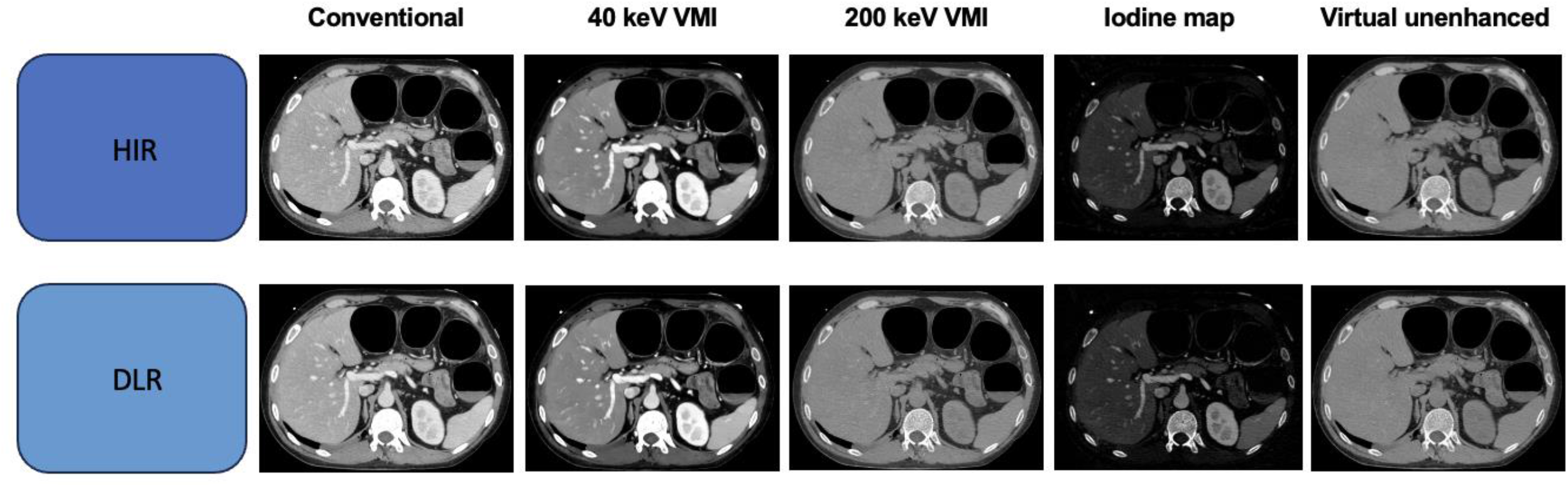
Example spectral output from the Philips CT 7500 scanner reconstructed with both the DLR examined in this manuscript and the standard HIR. All images are shown at W:350/L:40 except for 40 keV VMI, which is shown at W:756/L:216.

Quantitative improvements do not by themselves establish that interpreting physicians will accept the resulting images. Objective image-quality metrics correlate only imperfectly with radiologists’ preferences,^19^ so blinded reader-based assessment of diagnostic image quality remains necessary to validate a reconstruction algorithm for clinical use.^19,20^ For the spectral reconstruction, reader-based qualitative image quality and physician preference, evaluated across diverse anatomy and across both conventional and spectral image types, have not been established.

Here we present a post hoc secondary analysis of a multi-institution reader study comparing this spectral deep learning reconstruction with the current standard hybrid iterative reconstruction across head, chest, abdomen/pelvis, and cardiac CT, for both conventional and virtual monoenergetic images.

## Materials and Methods

### Study design

This study was a retrospective, post hoc secondary analysis of a multi-institutional, controlled, blinded reader study comparing two reconstruction methods (HIR and DLR) applied to identical raw CT data. The parent study included 147 CT examinations acquired and reconstructed across a range of technical parameters including standard and high-resolution acquisitions and both soft tissue and bone kernels. The present post hoc analysis used previously collected reader ratings from a subset of the original cohort that were acquired with standard resolution and reconstructed with thin sections and a soft tissue kernel. Restricting the analysis to these technical parameters allowed comparison to focus on DLR versus HIR.

The parent study was registered at ClinicalTrials.gov before enrollment of the first participant (NCT07108205) and was approved by the institutional review board of each participating institution (IRB Protocol # 857614 and 25-08-209-01). The requirements for written informed consent and Health Insurance Portability and Accountability Act (HIPAA) authorization were waived because of the retrospective use of deidentified data. The parent study was conducted at two urban academic healthcare systems and involved no changes to clinical workflow.

### Patient cohort

As part of the parent study, examinations were reviewed retrospectively in chronological order to ensure the availability of raw (projection-domain) CT data. Eligible examinations were head, chest, cardiac, and abdomen/pelvis CT studies performed on a dual-layer detector spectral CT system (CT 7500, Philips Healthcare) between July 16, 2025 and November 23, 2025. Inclusion criteria were age older than 22 years and a head, chest, cardiac, or abdomen/pelvis CT examination. The exclusion criterion was a non-diagnostic examination (e.g., substantial motion or streak artifact), as determined by the principal investigator. For each included examination, the raw data were de-identified and transferred to the manufacturer (Philips Healthcare) for reconstruction.

### Image reconstruction

The raw data comprised the high- and low-energy photon signals recorded by the two detector layers. For every examination, the data were reconstructed to generate a conventional and VMI result. Conventional images were produced by combining the two detector-layer signals into a single energy bin. Spectral images were reconstructed as VMIs (66 keV for head; 70 keV for chest, abdomen/pelvis, and cardiac). All examinations used in this post hoc secondary analysis were reconstructed with a soft tissue kernel and thin sections (3 mm for head; 2 mm for all other regions). Each conventional and each spectral dataset was reconstructed twice from the identical raw data: once with the current clinical-standard HIR (iDose^4^, Philips Healthcare) and once with DLR (Spectral Precise Image, Philips Healthcare). This yielded four reconstructions per examination: conventional HIR, conventional DLR, VMI HIR, and VMI DLR.

### Reader review and image-quality assessment

As part of the parent reader study, the reconstructed image sets were uploaded to a web-based image-review platform (Medidata Solutions, New York, NY), which provided each reader with an individual login, structured onboarding, and an integrated DICOM viewer with window/level adjustment, measurement tools, and series synchronization, as well as an audit trail of assigned examinations and recorded ratings. Reconstruction pairs were presented in randomized order, and readers were blinded to reconstruction method. For each assigned examination, readers directly compared the DLR and HIR reconstructions of the same images, separately for the conventional and the VMI images. For each comparison, readers recorded: (1) the overall image quality of each reconstruction on a 5-point Likert scale (1 = Poor image quality, nondiagnostic, unsatisfactory, 5 = Excellent image quality, fully diagnostic, satisfactory); (2) whether artifact was present in either series; (3) whether image texture was similar between the two reconstructions or superior in one. The body cohort included chest and abdomen/pelvis examinations. Each examination received two image quality scores, one for the primary region and one for the partially imaged region (for example, a CT abdomen/pelvis received both a score for the abdomen and for the partially imaged chest); image texture was scored once for the examination.

Each examination was reviewed independently by two readers. A total of 13 readers participated. All examinations were interpreted by U.S. board-certified radiologists or cardiologists (cardiac only), each with at least 2 years of post-training experience.

### Endpoints

The primary endpoint was overall image quality (Likert score), evaluated as noninferiority and superiority (assessed only when noninferiority was established) of DLR relative to HIR within each anatomic group and image type (conventional and VMI). Secondary endpoints were success rate (defined as the proportion of image pairs for which both readers assigned DLR a score that was at least as high as the corresponding HIR score and was 3 or higher), reader-assessed image texture, the frequency and clinical impact of artifacts, and interobserver agreement.

### Sample-size determination and analytic cohort

Sample size considerations for the parent study were based on detecting a mean between-reconstruction difference in Likert score of 0.4, assuming a standard deviation of 1.1, a noninferiority margin of −0.10, 80% power, and a one-sided significance level of 0.025. The sample size for this post hoc secondary analysis was determined by the number of examinations in the parent reader study that met the technical criteria of standard resolution acquisition, thin-section reconstruction and soft tissue kernel, which included 15 head, 22 body, and 22 cardiac CT examinations. Body examinations contributed both chest and abdomen anatomic-region assessments, yielding 81 scored anatomic regions: 15 head, 22 chest, 22 abdomen, and 22 cardiac. The post hoc power was 0.35 for 15 examinations and 0.50 for 22 examinations.

### Statistical analysis

Overall image-quality scores were summarized as least-squares (LS) mean Likert scores for each reconstruction method within each anatomic group. Head, cardiac, chest, and abdomen/pelvis examinations were analyzed separately. LS means were estimated from a model that accounted for multiple readers per examination with reconstruction method as a fixed effect. Noninferiority of DLR to HIR was concluded, at a one-sided 2.5% significance level, if the lower bound of the two-sided 95% confidence interval for the difference in LS means (DLR − HIR) exceeded the pre-specified noninferiority margin of −0.10. Where noninferiority was established, superiority of DLR was tested from the same model at a two-sided significance level of 2.5%. Success rate was defined as the proportion of image pairs for which both readers assigned DLR a score that was at least as high as the corresponding HIR score and was 3 or higher. Reported p values correspond to superiority testing. Interobserver agreement between the two readers of each image set was assessed with the weighted Cohen κ across the full 5-point scale, separately for anatomic group and image type (conventional or VMI), for both reconstruction methods (HIR or DLR). The pre-specified agreement endpoint was a weighted κ greater than 0.80 for DLR. As this endpoint was not met, an exploratory analysis was performed, in which Likert scores were pooled into three categories—Poor (1–2), Fair (3), and Good (4–5)—and the corresponding percent agreement (agreement rate), with 95% Wald-type confidence intervals, was calculated. Analyses were performed using SAS (SAS® Life Science Analytics Framework, Release: 5.4.1b, SAS Institute Inc., Cary, NC, USA).

## Results

### Cohort

A total of 59 CT examinations from 56 patients met the criteria for the present secondary analysis. No examinations were subsequently excluded (Figure 2). Three patients underwent both a chest CT and an abdomen/pelvis CT as separate acquisitions; each acquisition was treated as a separate examination in the image-quality analysis. Examination-level demographic and scan characteristics are summarized in Tables 1 and 2. Across 81 anatomic regions available for scoring (15 head, 22 chest, 22 abdomen, 22 cardiac), two image types (conventional and VMI), two reconstruction methods (DLR and HIR), and two readers, 648 individual Likert scores were possible. Three scores were missing, leaving 645 available scores. Because paired analysis required both DLR and HIR scores, 321 paired comparisons were available comprising 642 Likert scores.

**Table 1.** Demographic characteristics. Data are presented at the examination level. Note: 59 examinations were obtained from 56 patients; three patients each contributed separate chest and abdomen/pelvis examinations.

|  | Head (N=15) | Body (N=22) | Cardiac (N=22) | Total (N=59) |
| --- | --- | --- | --- | --- |
| **Sex [n(%)]** |  |  |  |  |
| Female | 7 (46.7%) | 11 (50.0%) | 13 (59.1%) | 31 (52.5%) |
| Male | 8 (53.3%) | 11 (50.0%) | 9 (40.9%) | 28 (47.5%) |
| **Age (years)** |  |  |  |  |
| Mean (SD) | 63.4 (21.15) | 62.4 (16.66) | 62.5 (13.87) | 62.7 (16.68) |
| Median (IQR) | 72.0 (37.0-80.0) | 66.0 (50.0-71.0) | 64.0 (56.0-69.0) | 66.0 (50.0-76.0) |
| **Race [n (%)]** |  |  |  |  |
| Asian | 2 (13.3%) | 2 (9.1%) | 1 (4.8%) | 5 (8.6%) |
| Black or African American | 3 (20.0%) | 5 (22.7%) | 6 (28.6%) | 14 (24.1%) |
| White | 10 (66.6%) | 9 (40.9%) | 0 | 19 (32.8%) |
| Multiple | 0 (0.0%) | 6 (27.3%) | 14 (66.7%) | 20 (34.5%) |
| Missing | 0 | 0 | 1 | 1 |
| **Height (cm)** |  |  |  |  |
| Mean (SD) | 168.8 (15.79) | 169.8 (12.58) | 163.9 (12.03) | 167.4 (13.31) |
| Median (IQR) | 165.1 (154.9-180.3) | 170.2 (162.6-177.8) | 165.1 (157.5-170.2) | 165.1 (157.5-172.7) |
| **Weight (kg)** |  |  |  |  |
| Mean (SD) | 76.6 (26.66) | 74.0 (17.89) | 77.6 (15.80) | 76.0 (19.50) |
| Median (IQR) | 67.1 (54.9-99.3) | 71.2 (58.5-89.8) | 76.5 (68.0-81.2) | 74.8 (61.2-88.9) |
| **BMI (kg/m<sup>2</sup>)** |  |  |  |  |
| Mean (SD) | 26.9 (8.10) | 25.6 (5.28) | 28.8 (4.09) | 27.1 (5.83) |
| Median (IQR) | 25.2 (17.9-34.9) | 24.6 (21.5-30.4) | 28.1 (25.5-30.9) | 26.2 (22.9-30.9) |

**Table 2.** CT imaging details.

|  | Head (N=15) | Body (N=22) | Cardiac (N=22) | Total (N=59) |
| --- | --- | --- | --- | --- |
| **kVp [n]** |  |  |  |  |
| 100 | 0 (0%) | 2 (9.1%) | 11 (50.0%) | 13 (22.0%) |
| 120 | 15 (100%) | 20 (90.9%) | 11 (50.0%) | 46 (78.0%) |
| **mAs** |  |  |  |  |
| Mean (SD) | 315.5 (24.65) | 153.0 (89.23) | 316.4 (66.27) | 255.3 (104.61) |
| Median (IQR) | 317.0 (299.0-332.0) | 147.0 (112.0-175.0) | 329.0 (261.0-360.0) | 276.0 (155.0-338.0) |
| **Rotation time** |  |  |  |  |
| Mean (SD) | 0.5 (0.07) | 0.5 (0.17) | 0.3 (0.00) | 0.4 (0.15) |
| Median (IQR) | 0.5 (0.5-0.5) | 0.4 (0.4-0.5) | 0.3 (0.3-0.3) | 0.4 (0.3-0.5) |
| **Contrast** |  |  |  |  |
| No | 15 (100%) | 8 (36.4%) | 0 (0%) | 23 (39.0%) |
| Yes | 0 (0%) | 14 (63.6%) | 22 (100%) | 36 (61.0%) |
| **Pitch** |  |  |  |  |
| Mean (SD) | 0.4 (0.00) | 1.1 (0.05) | 0.2 (0.00) | 0.7 (0.43) |
| Median (IQR) | 0.4 (0.4-0.4) | 1.2 (1.2-1.2) | 0.2 (0.2-0.2) | 0.4 (0.4-1.2) |
| **Dose (mGy)** |  |  |  |  |
| Mean (SD) | 57.3 (15.56) | 12.6 (7.38) | 22.3 (6.19) | 27.6 (20.41) |
| Median (IQR) | 58.2 (55.5-68.1) | 11.0 (8.7-14.4) | 21.0 (18.1-23.5) | 20.4 (13.7-36.5) |

**Figure 2.**
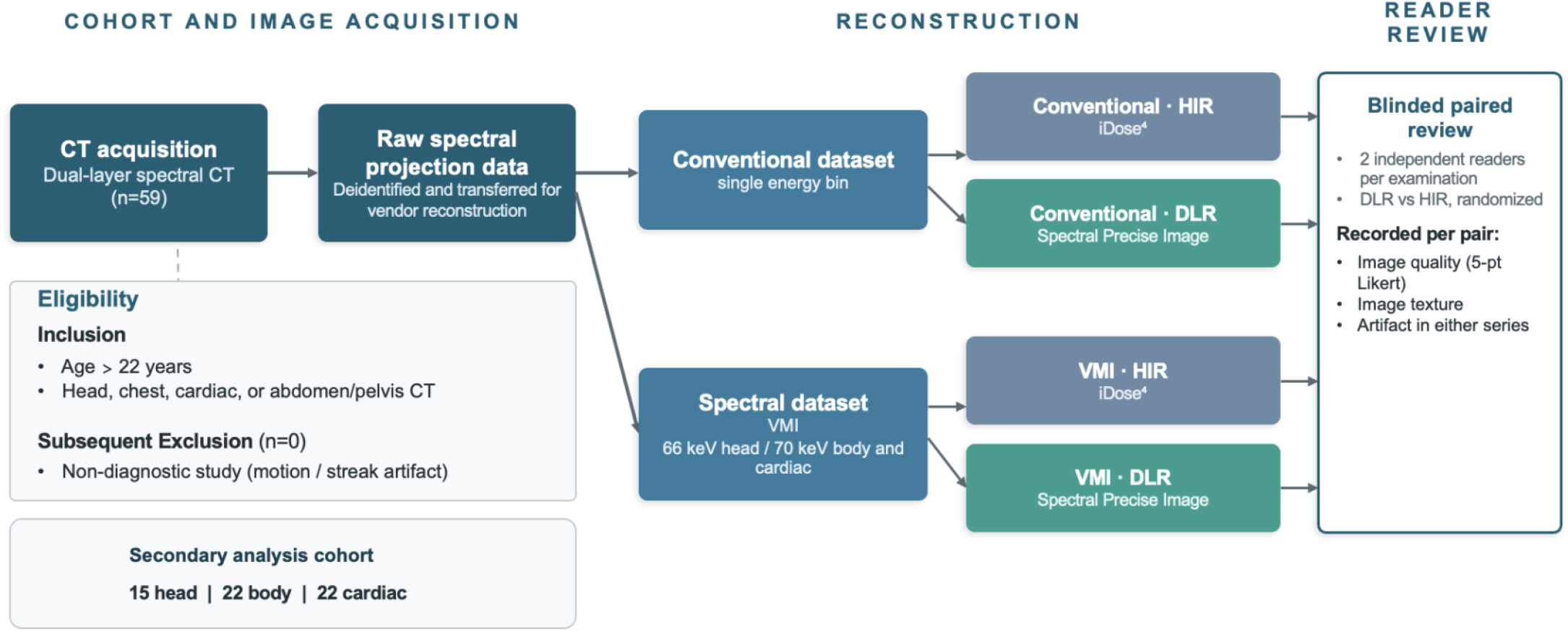
Study workflow of the post hoc secondary analysis. Previously collected ratings from 59 standard-resolution examinations reconstructed with thin sections and a soft-tissue filter were selected from the 147-examination parent reader study.

### Image quality

DLR was both noninferior and superior to HIR across head, abdomen and cardiac groups for both conventional and VMI, with p values ranging from <.0001 to .0058 (Table 3). DLR was noninferior but not superior for conventional chest images. Noninferiority was not established for chest VMI, although mean scores numerically favored DLR. Success rates for conventional images were 100% (95% CI 78-100%), 96% (71-99%), 91% (66-98%) and 96% (71-99%) for head, abdomen, chest and cardiac respectively; corresponding VMI success rates were 100% (78-100%), 100% (85-100%), 96% (71-99%) and 95% (70-99%). See Figure 3 and Supplemental Figures 1 and 2 for representative paired readings.

**Table 3.**
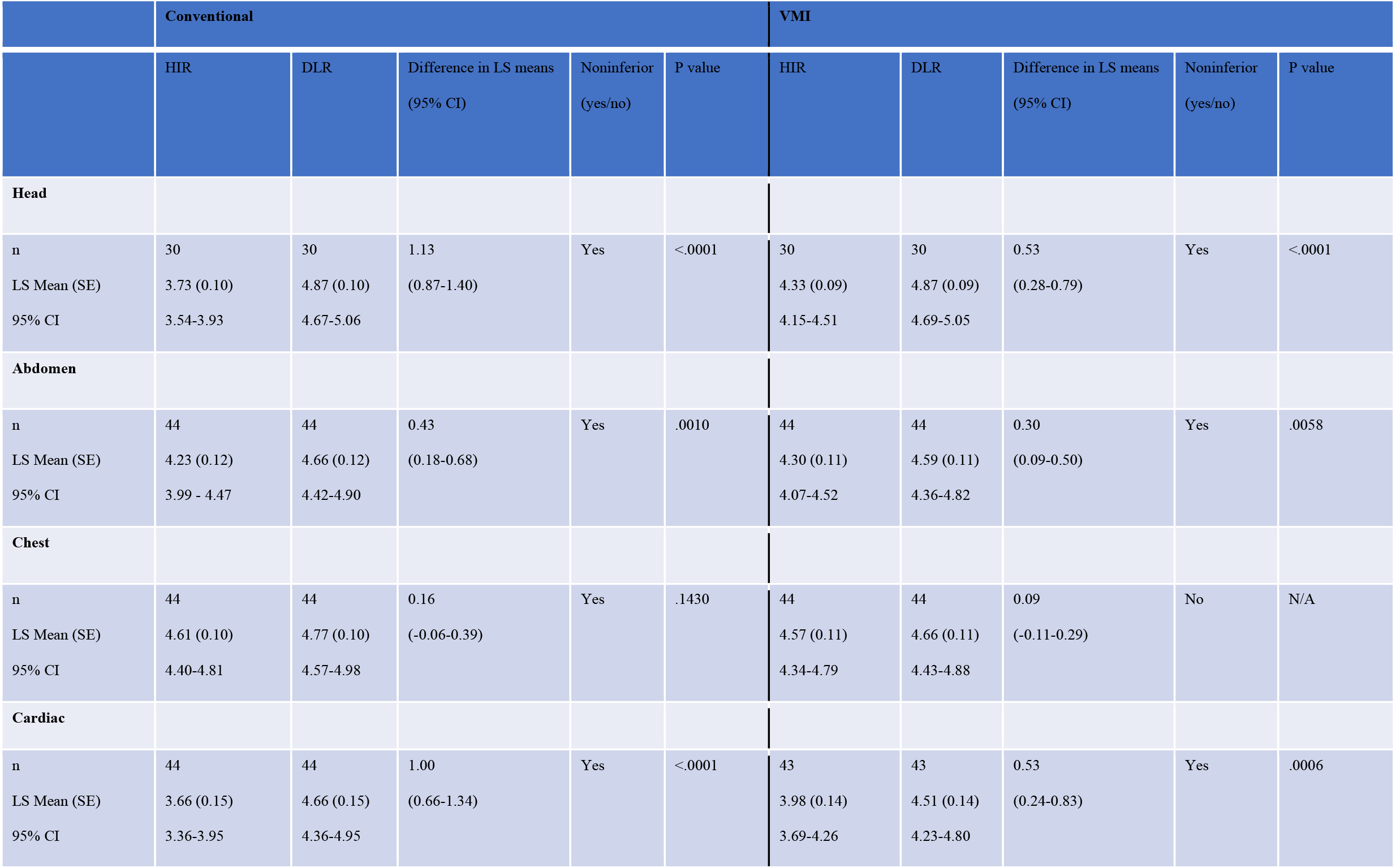
LS mean Likert scores of HIR compared to DLR for both conventional and VMI across all anatomic groups. A 70 keV VMI was used for abdomen, chest and cardiac CT. 66 keV VMI was used for Head CT. Noninferiority is met when the lower bound of the two-sided 95% confidence interval for the difference in LS means is above -0.1. P values correspond to superiority testing when noninferiority was established; superiority was not formally tested for chest VMI. n=number of reads.

|  | Conventional |  |  |  |  | VMI |  |  |  |  |
| --- | --- | --- | --- | --- | --- | --- | --- | --- | --- | --- |
|  | HIR | DLR | Difference in LS means<br>(95% CI) | Noninferior<br>(yes/no) | P value | HIR | DLR | Difference in LS means<br>(95% CI) | Noninferior<br>(yes/no) | P value |
| **Head** |  |  |  |  |  |  |  |  |  |  |
| n | 30 | 30 | 1.13 | Yes | <.0001 | 30 | 30 | 0.53 | Yes | <.0001 |
| LS Mean (SE) | 3.73 (0.10) | 4.87 (0.10) | (0.87-1.40) |  |  | 4.33 (0.09) | 4.87 (0.09) | (0.28-0.79) |  |  |
| 95% CI | 3.54-3.93 | 4.67-5.06 |  |  |  | 4.15-4.51 | 4.69-5.05 |  |  |  |
| **Abdomen** |  |  |  |  |  |  |  |  |  |  |
| n | 44 | 44 | 0.43 | Yes | .0010 | 44 | 44 | 0.30 | Yes | .0058 |
| LS Mean (SE) | 4.23 (0.12) | 4.66 (0.12) | (0.18-0.68) |  |  | 4.30 (0.11) | 4.59 (0.11) | (0.09-0.50) |  |  |
| 95% CI | 3.99 - 4.47 | 4.42-4.90 |  |  |  | 4.07-4.52 | 4.36-4.82 |  |  |  |
| **Chest** |  |  |  |  |  |  |  |  |  |  |
| n | 44 | 44 | 0.16 | Yes | .1430 | 44 | 44 | 0.09 | No | N/A |
| LS Mean (SE) | 4.61 (0.10) | 4.77 (0.10) | (-0.06-0.39) |  |  | 4.57 (0.11) | 4.66 (0.11) | (-0.11-0.29) |  |  |
| 95% CI | 4.40-4.81 | 4.57-4.98 |  |  |  | 4.34-4.79 | 4.43-4.88 |  |  |  |
| **Cardiac** |  |  |  |  |  |  |  |  |  |  |
| n | 44 | 44 | 1.00 | Yes | <.0001 | 43 | 43 | 0.53 | Yes | .0006 |
| LS Mean (SE) | 3.66 (0.15) | 4.66 (0.15) | (0.66-1.34) |  |  | 3.98 (0.14) | 4.51 (0.14) | (0.24-0.83) |  |  |
| 95% CI | 3.36-3.95 | 4.36-4.95 |  |  |  | 3.69-4.26 | 4.23-4.80 |  |  |  |

**Figure 3.**
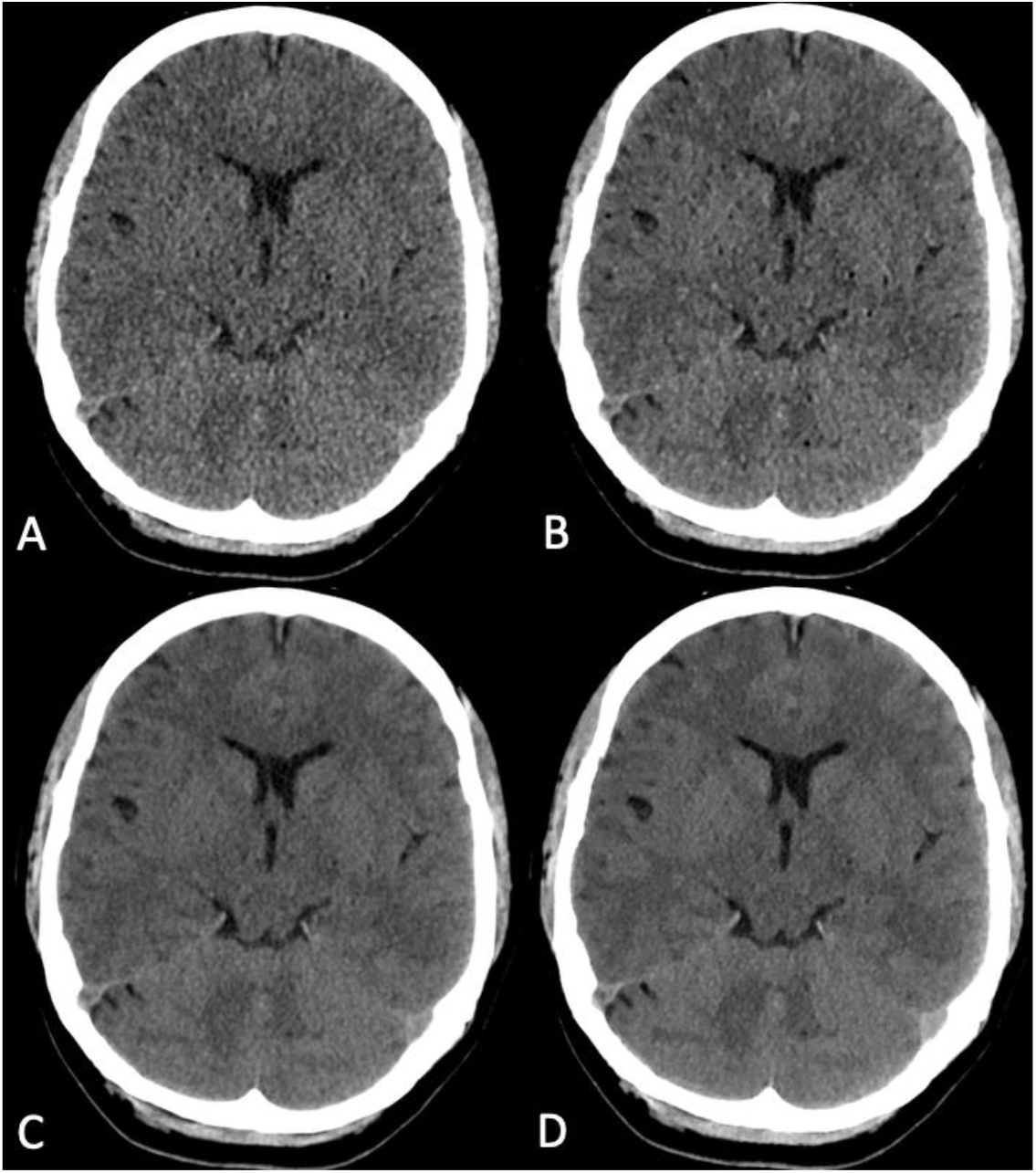
Axial CT of the head. All 4 images were reconstructed using a soft-tissue filter and are presented using the same windowing (W:100 L:50). A. Conventional dataset reconstructed with HIR. Reader 1 score: 4, Reader 2 score: 3. B. VMI (66 keV) dataset reconstructed with HIR. Reader 1 score: 4, Reader 2 score: 3. C. Conventional dataset reconstructed with DLR. Reader 1 score: 5, Reader 2 score: 5. D. VMI (66 keV) dataset reconstructed with DLR. Reader 1 score: 5, Reader 2 score: 5.

### Interobserver agreement

The weighted Cohen’s kappa values across the full 5-point scale were low (ranging between 0.04 and 0.23) and did not meet the endpoint of 0.80.

In post hoc analysis, agreement rates for HIR were 28.2% (95% CI 18.0%-41.2%) and 37.5% (27.4%-48.8%) for conventional and VMI images respectively. Agreement rates for DLR were 67.4% (54.7%-78.0%) and 62.2% (48.4%-74.2%) for conventional and VMI images respectively.

When comparing across anatomic groups, agreement rates for HIR were 33.3% (17.9%-53.4%), 38.5% (23.4%-56.0%), 44.0% (25.2%-64.6%), and 18.4% (8.7%-34.9%) for head, abdomen, chest and cardiac respectively. Agreement rates for DLR were 74.6% (49.5%-89.8%), 65.8% (42.0%-83.7%), 60.7% (36.7%-80.4%), 64.0% (43.2%-80.6%) for head, abdomen, chest and cardiac respectively.

Therefore, agreement rates were higher in DLR across anatomic groups for both conventional and VMI images.

### Image Texture

DLR was rated as having similar or better image texture in 100% conventional and VMI head and body CT reads, 100% of conventional cardiac CT reads and 95.3% of VMI cardiac CT reads (Table 4).

**Table 4.**
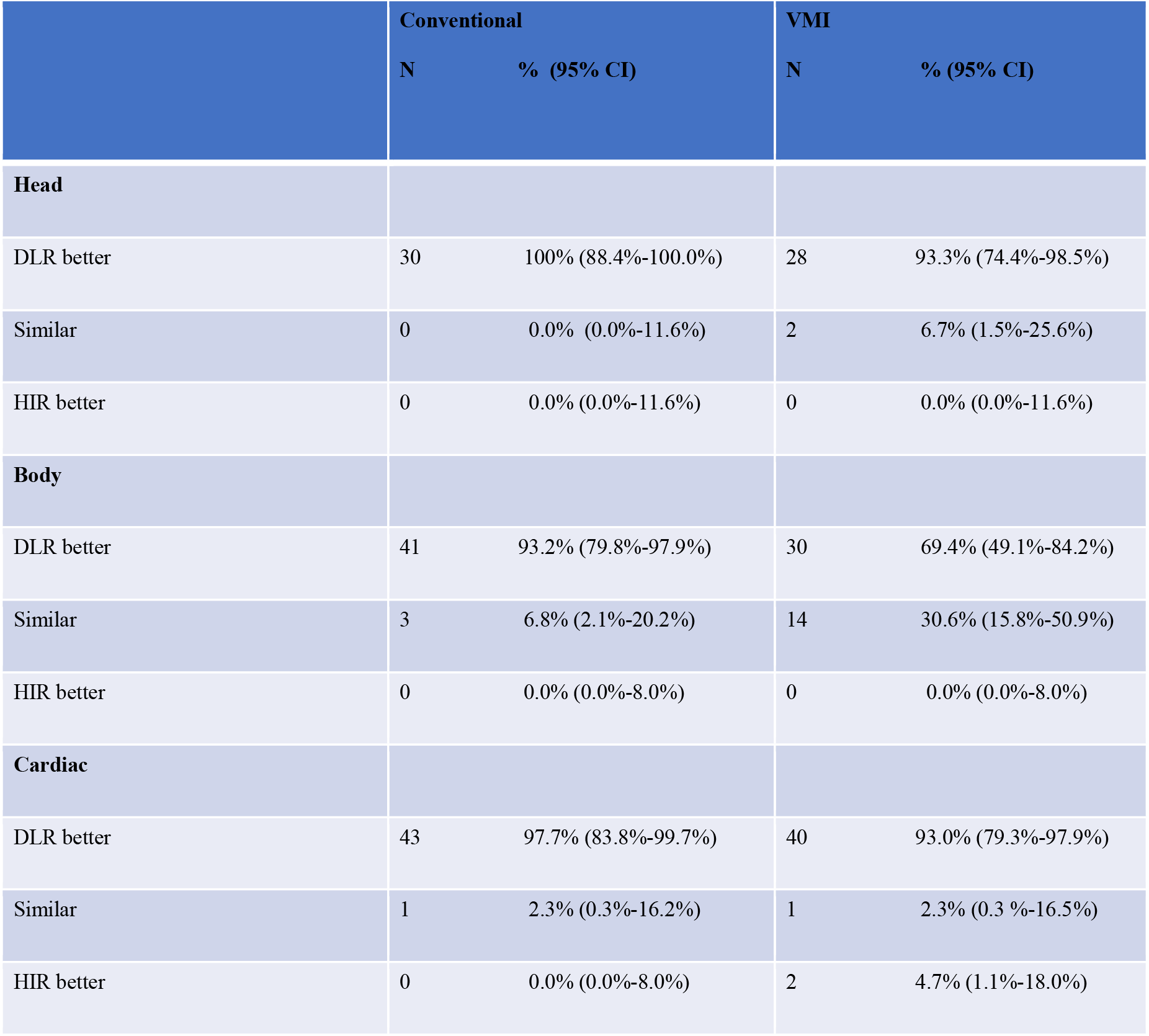
Reader-assessed image texture preference by reconstruction method. DLR image texture was rated similar or superior to HIR in nearly all reads across conventional images and VMI. N=the number of individual reads.

|  | Conventional |  | VMI |  |
| --- | --- | --- | --- | --- |
|  | N | % (95% CI) | N | % (95% CI) |
| **Head** |  |  |  |  |
| DLR better | 30 | 100% (88.4%-100.0%) | 28 | 93.3% (74.4%-98.5%) |
| Similar | 0 | 0.0% (0.0%-11.6%) | 2 | 6.7% (1.5%-25.6%) |
| HIR better | 0 | 0.0% (0.0%-11.6%) | 0 | 0.0% (0.0%-11.6%) |
| **Body** |  |  |  |  |
| DLR better | 41 | 93.2% (79.8%-97.9%) | 30 | 69.4% (49.1%-84.2%) |
| Similar | 3 | 6.8% (2.1%-20.2%) | 14 | 30.6% (15.8%-50.9%) |
| HIR better | 0 | 0.0% (0.0%-8.0%) | 0 | 0.0% (0.0%-8.0%) |
| **Cardiac** |  |  |  |  |
| DLR better | 43 | 97.7% (83.8%-99.7%) | 40 | 93.0% (79.3%-97.9%) |
| Similar | 1 | 2.3% (0.3%-16.2%) | 1 | 2.3% (0.3 %-16.5%) |
| HIR better | 0 | 0.0% (0.0%-8.0%) | 2 | 4.7% (1.1%-18.0%) |

### Artifacts

The most common artifact was caused by the presence of metal. It was reported by both readers on both image types, conventional and VMI, in 3/15 head CT examinations and 2/22 body CT examinations. In 1/15 head CT examination and 5/22 body CT examinations, metal was reported by only one reader on both image types. In an additional 1/22 body CT examination, metal was reported by one reader on the conventional images only. These artifacts were not observed to affect clinical assessment. No metal artifacts were reported in the cardiac CT examinations.

## Discussion

In this retrospective, post hoc secondary analysis of a multi-institution reader study, spectral DLR produced image quality that was noninferior to HIR for most evaluated anatomic regions and image types, and was superior for head, abdomen, and cardiac regions for both conventional images and VMI. Chest VMI was the only setting in which noninferiority was not established, although mean scores numerically favored DLR. To our knowledge, this is the first multi-institution, blinded reader study to validate the spectral implementation of this algorithm across diverse anatomy and both conventional and VMI image types, extending prior evidence that has been largely phantom-based, single-center, or limited to the quantitative performance of the algorithm.

The clinical relevance of these findings follows directly from how spectral images are generated. Because material decomposition is mathematically ill-posed and amplifies noise, raw-data noise propagates into every spectral-derived image; a reconstruction that reduces this noise while preserving image texture is therefore expected to benefit conventional and spectral images simultaneously.^12,13^ Our reader data bear this out: DLR was preferred not only on conventional images but also on VMI in three of the four anatomic regions, indicating that the texture-preserving noise reduction radiologists value on conventional images carries over to the VMI images derived from the same acquisition.

In the chest, DLR retained a numerical advantage over HIR for both conventional (4.77 vs 4.61) and VMI (4.66 vs 4.57), although the difference did not reach the prespecified thresholds for superiority or, for VMI, noninferiority. This pattern likely reflects a ceiling effect: HIR achieved its highest scores of any region in the chest, leaving little room on the 5-point scale for either reconstruction to separate.^21^ The chest also has high intrinsic contrast among air, fat, and vessels, so image quality there is less limited by noise—and noise reduction is precisely where DLR contributes most, as reflected in the larger gains observed in the lower-contrast brain, liver, and cardiac examinations. DLR was therefore at no practical disadvantage in the chest, and future work with larger chest cohorts and a wider range of protocols is likely to be better positioned to resolve the small residual differences in this already high-performing region.

Inter-reader agreement, measured by the weighted Cohen κ, was low for both reconstructions. This most likely reflects the well-described κ paradox: when ratings cluster in the upper categories, as they did here, κ is driven downward even when raters agree in absolute terms, so these low values should not be read as poor reliability.^22,23^ When agreement was instead expressed as the proportion of image sets for which both readers assigned the same pooled category (Poor [1-2], Fair [3], or Good [4-5]), agreement was consistently higher for DLR than for HIR across anatomic groups and for both conventional images and VMI (for example, 67.4% vs 28.2% agreement on conventional images). This higher agreement rate, considered together with the higher mean Likert scores for DLR, suggests DLR more reliably produced images that radiologists rated favorably.

Clinicians likely favored DLR because the images were perceived as having reduced image noise while preserving natural image texture. DLR was rated similar or superior in texture across nearly all evaluations—100% of conventional and VMI head and body reads, 100% of conventional cardiac reads, and 95.3% of cardiac VMI reads—suggesting that the improvement in perceived image quality was not accompanied by an over-smoothed appearance. Metal was reported on several examinations; however no reported metal artifact affected clinical assessment.

The principal strengths of this study are its multi-institution design and the large number of examinations from a diverse patient demographic reviewed under blinded, randomized, paired conditions with two independent readers per examination. Several limitations should be acknowledged. This was a post hoc analysis only evaluating examinations acquired at standard resolution and reconstructed with a soft tissue kernel and thin sections to isolate the effect of DLR versus HIR. This restriction reduced technical heterogeneity but may introduce selection bias and limits generalizability to high-resolution acquisitions and bone kernel reconstructions evaluated in the parent study. The parent reader study included other acquisition resolutions, kernels, and slice thicknesses but their comparative performance was not evaluated in the present analysis. The assessment was qualitative and, despite explicitly defined scoring criteria, retained an element of subjectivity, and the per-group sample sizes—while sufficient to demonstrate the effects reported here—were modest and of limited power to detect smaller differences. Finally, this study evaluated perceived image quality rather than diagnostic accuracy; whether the reconstruction affects lesion detection or characterization must be established in future studies.^24,25^

In conclusion, this spectral deep learning reconstruction produced image quality that matched or exceeded standard hybrid iterative reconstruction across most anatomic regions and image types, including conventional and virtual monoenergetic images, with more consistent reader agreement and preserved image texture.

## Data Availability

All data produced in the present study are available upon reasonable request to the authors

## Acknowledgements

Microsoft 365 Copilot (Microsoft; GPT-5 reasoning model; accessed August 2026) was used for editing, grammar, clarity and manuscript organization. It was not used to generate or analyze study data, perform statistical analyses, or create clinical images. All AI-assisted content was reviewed and approved by the authors.

We acknowledge support from Philips Healthcare.

