## Supplemental Figures 1 and 2 for "Deep Learning Reconstruction versus Hybrid Iterative Reconstruction for Spectral CT: A Multi-Institutional Image Quality Assessment"

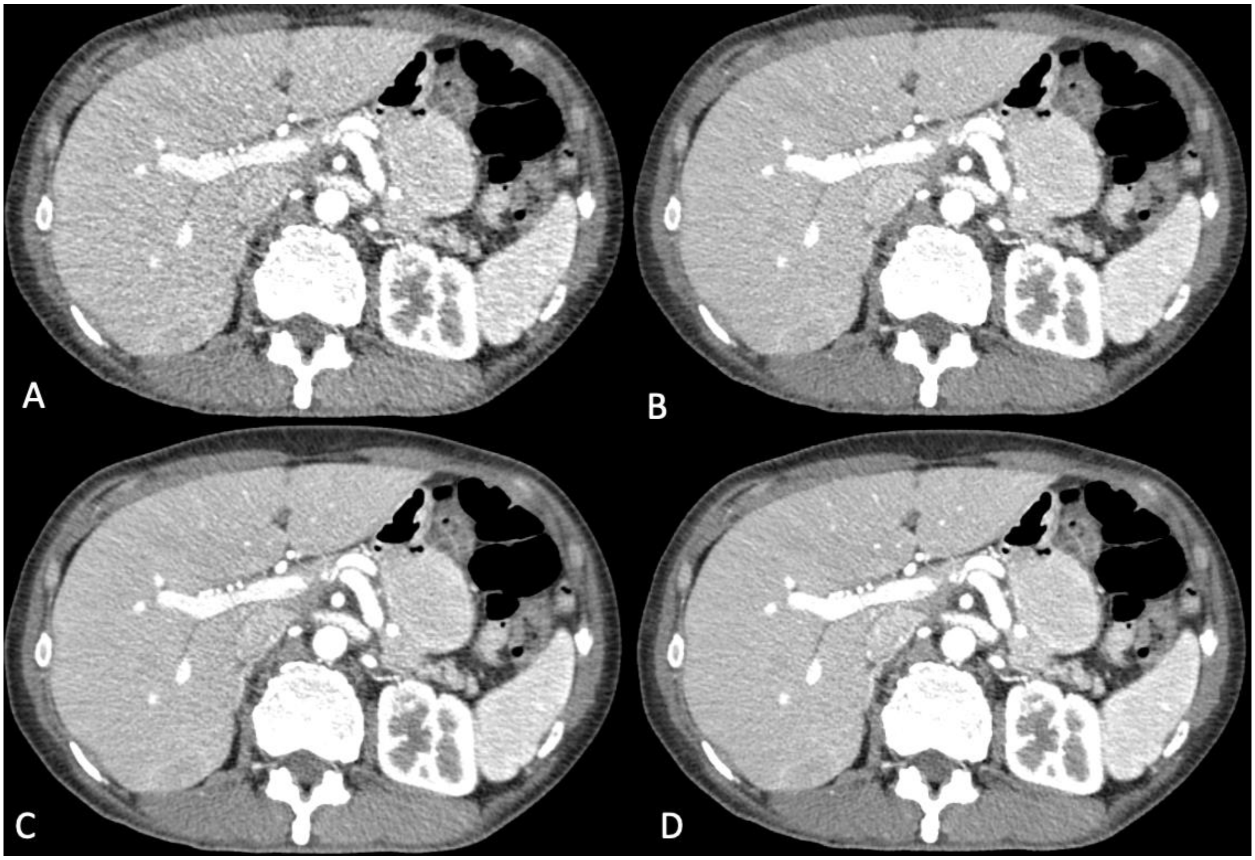


Supplemental Figure 1. Contrast enhanced CT through the abdomen. All 4 images were reconstructed using a soft-tissue filter and are presented using the same windowing (W:350 L:40). A. Conventional dataset reconstructed with HIR. Reader 1 score: 5, Reader 2 score: 3. B. VMI (70 keV) dataset reconstructed with HIR. Reader 1 score: 5, Reader 2 score: 3. C. Conventional dataset reconstructed with DLR. Reader 1 score: 5, Reader 2 score: 4. D. VMI (70 keV) dataset reconstructed with DLR. Reader 1 score: 5, Reader 2 score: 4.

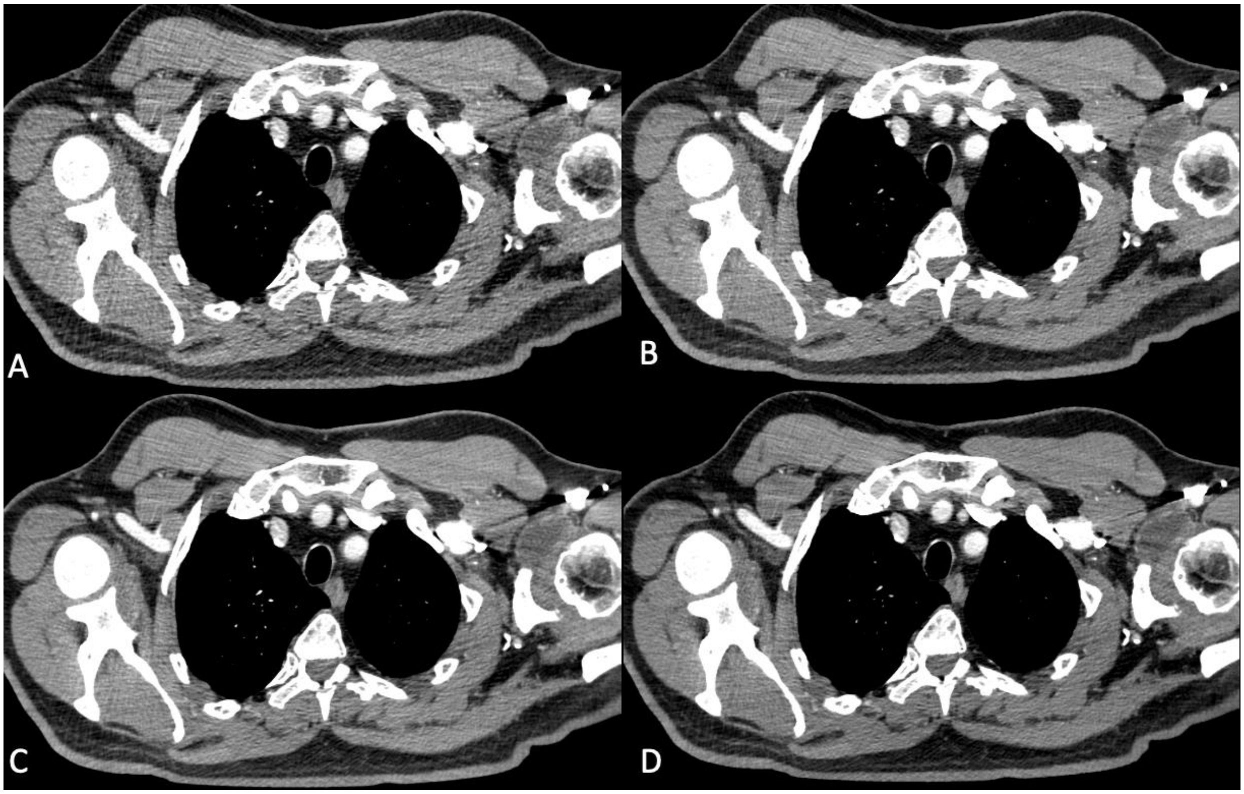


Supplemental Figure 2. Contrast enhanced CT through the chest. All 4 images were reconstructed using a soft-tissue filter and are presented using the same windowing (W:350 L:40). A. Conventional dataset reconstructed with HIR. Reader 1 score: 5, Reader 2 score: 4. B. VMI (70 keV) dataset reconstructed with HIR. Reader 1 score: 5, Reader 2 score: 4.  C. Conventional dataset reconstructed with DLR. Reader 1 score: 5, Reader 2 score: 4. D. VMI (70 keV) dataset reconstructed with DLR. Reader 1 score: 5, Reader 2 score: 4.
